# Sex-specific longitudinal changes in resting heart rate and all-cause heart failure: insights from the HUNT Study

**DOI:** 10.1101/2025.11.06.25339723

**Authors:** Linda MS. Hansen, Sumaya SH. Jui, Tonje B. Braaten, Håvard Dalen, Lars B. Forr-Garnvik, Trine Karlsen

**Author notes:** Corresponding author: Name: Trine Karlsen, Address: Universitetsalléen 11, 8026 Bodø, Norway.

## Abstract

**Background:** Investigation of sex-specific associations between longitudinal resting heart rate (RHR) and new-onset heart failure (HF) using both change in RHR and RHR trajectories.

**Methods:** Participants from the Trøndelag Health Study attending two or three surveys between 1995-2019 were included. We investigated the association between new-onset HF and RHR, using RHR categories according to the mean baseline RHR standard deviation (12 bpm), continuous RHR in restricted cubic splines (n=47712, mean 12-year follow-up), and latent class trajectory models (n=47162, mean 7-year follow-up). Cox regression was used to estimate adjusted hazard ratios (HR) and 95% confidence intervals (95% CI).

**Results:** During follow-up, 2880 of the 47712 participants developed HF. The HF incidence rate was lower in women than men (4.27 vs. 5.68 per 1000 person-years, ratio (95% CI) 0.67 (0.57-0.77). Baseline RHR was 74 bpm in women and 70 bpm in men, and 74% maintained their RHR (±12 bpm) from baseline to their second attendance (mean change -2±12 bpm). Each 10-bpm higher RHR was associated with higher risk of HF for both women and men with HR (95% CI) 1.15 (1.03-1.28) and 1.09 (1.00-1.20), respectively. Participants with a high RHR trajectory had higher risk of HF compared to the low RHR-trajectories with HR (95% CI) 1.43 (1.14-1.79) for women and 1.41 (1.16-1.72) for men.

**Conclusion:** All-cause HF was similarly associated with increased RHR and a high RHR trajectory for women and men. Estimating HF risk using RHR trajectories provided stronger associations between RHR and HF compared to using a single RHR measurement.

**Lay summary:** We investigated sex-specific associations between longitudinal resting heart rate (RHR) and all-cause new-onset of heart failure for 47 712 participants from the Trøndelag Health Study, using RHR categories, continuous RHR in restricted cubic splines, and latent class trajectory models.

- Each 10-bpm increase in RHR was associated with a similar higher risk of heart failure for both women and men, with 15% higher risk for women and 9% higher for men
- Participants with a high RHR trajectory had 41-43% increased risk of heart failure compared to the low RHR-trajectory
- Long term RHR measurements can reveal changes in HF risk profiles that might not be detectable with a single RHR measurement and highlight individuals with a high RHR who should be further evaluated

Graphical abstract.
Sex-specific associations between longitudinal resting heart rate (RHR) and heart failure (HF) from The Trøndelag Health Study, 1995-2019. In fully adjusted analyses, women and men with an increase in RHR or a high RHR trajectory had a similar risk of developing all-cause new-onset HF.

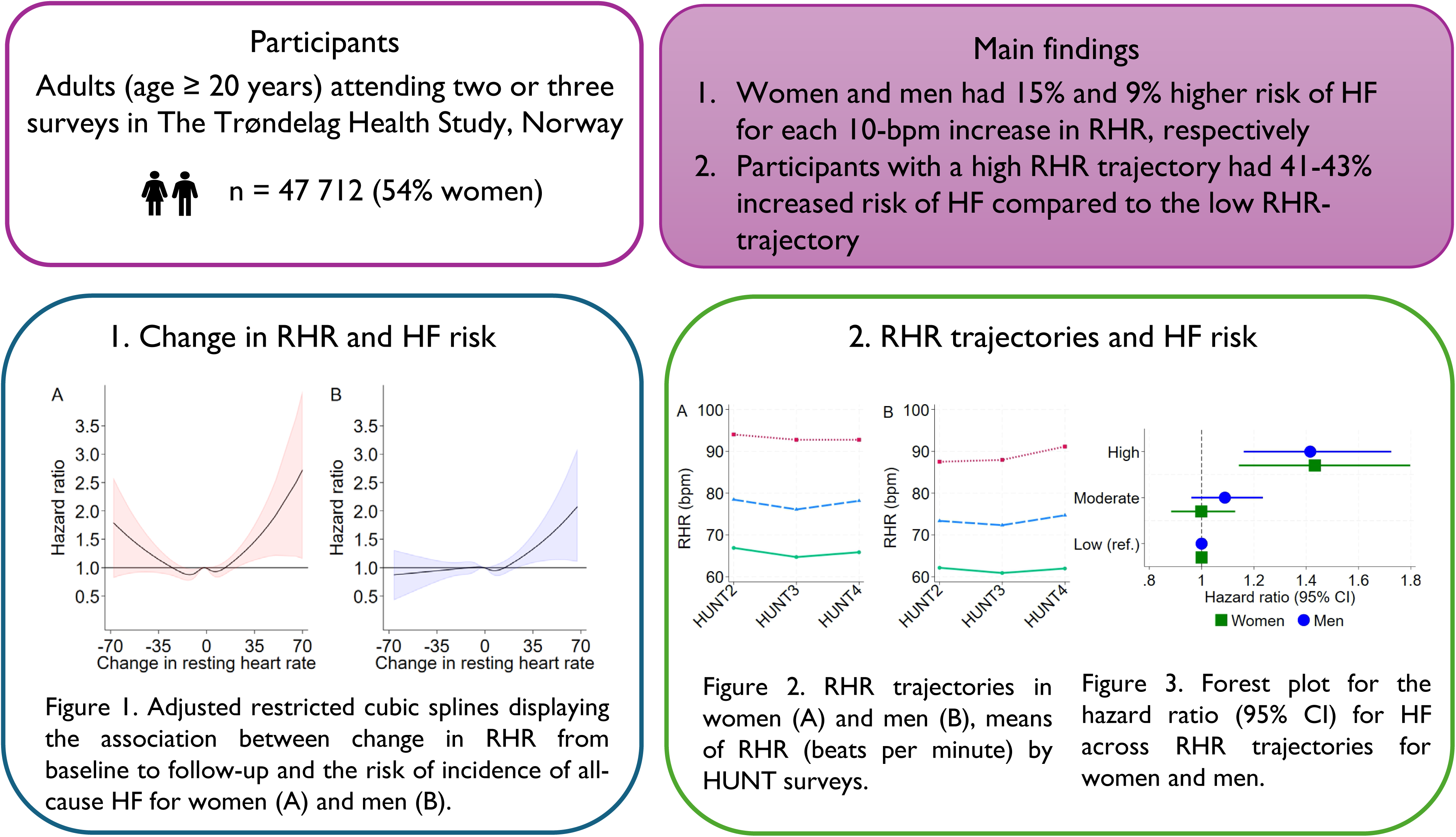

## Introduction

Heart failure (HF) affected around 64 million people globally in 2017^1^, with a similar lifetime risk of HF for European women and men after the age of 75 years^2^. Women generally develop HF 4-5 years later than men^2^, and risk factors like diabetes mellitus, hypertension, smoking, and obesity may impact all-cause HF more in women than men^3^. Additionally, hormone and immune responses, pregnancy complications and menopause might be specific risk factors for HF in women^4^. Although sex differences in HF subtypes and risk factors are acknowledged in preventive guidelines^5^, it is unclear if there are sex differences in potential prognostic markers like change in resting heart rate (RHR).

RHR is an easy non-invasive cardiac parameter previously associated with both risk factors for HF and its incidence^6,7^. RHR is generally 3-7 bpm higher for women than men and is also affected by age, genes, and lifestyle factors like physical activity and smoking^8^. A normal RHR (e.g. 60-84 bpm) reflects a physically active lifestyle, and a high RHR (e.g. ≥ 85 bpm) may imply the presence of risk factors for all-cause HF (e.g., higher age, hypertension, physical inactivity, smoking)^7^. In pooled sex analyses in two meta-analyses on data from cross-sectional studies, each 10-bpm higher RHR was associated with 18-19% higher relative risk of incident HF^6,9^. In a sex-specific analyses, men had 14% higher relative risk of HF for each 10-bmp higher RHR, with no such association in women^9^. A single measurement of RHR can reflect physiological and environmental risk factors for HF, but it is currently unknown if longitudinal RHR may better reflect long-term changes in cardiovascular physiology or pathophysiology^7^.

Previous studies have shown that changes in RHR are associated with all-cause mortality and ischemic heart disease^10–12^. Studies who investigated the associations between longitudinal RHR and HF did not exclude participants with pre-existing HF^13–15^ or typically had short time observations, with 3-5 years between RHR measurements^16,17^. One study reported higher risk of all-cause incident HF for women than men when comparing a high RHR trajectory (> 72 bpm) to a low RHR trajectory (< 60 bpm)^18^. To our knowledge, no previous studies have investigated the longitudinal relationship between RHR and HF using an integrated approach that combines categorical, continuous, and trajectory-based analyses. Moreover, long-term research on RHR trajectories and incidence HF remains scarce within European populations. We therefore investigated the sex-specific hazard ratio between longitudinal RHR and all-cause incidence of HF in a Norwegian adult cohort from the Trøndelag Health Study (HUNT) using both changes in RHR and RHR trajectories.

## Methods

### Study design and population

The Trøndelag Health Study (HUNT) cohort study invited all adult residents in the Nord-Trøndelag County, Norway, to participate in four study surveys between 1984-2019^19^. Data from 1995-97 (HUNT2), 2006-08 (HUNT3), and 2017-19 (HUNT4) were used in this study. Briefly, the 96 436 individuals participating in HUNT2, HUNT3 and/or HUNT4 were between 20-104 years old and 53% were women^19,20^. The participation rates were 70% in HUNT2 and 54% in HUNT3 and HUNT4^19^. Details of the HUNT study design and population are provided elsewhere^19,20^. We excluded 47880 participants (52% women) with less than two RHR measurements (50% of participants from HUNT2-4), and 831 participants (43% women) diagnosed with all-cause HF before the second RHR measurement from the categorical and continuous analyses (Figure 1). For the RHR trajectory analysis, we further excluded 550 participants (38% women) with three RHR measurements and all-cause HF diagnosed before HUNT4. Overall, we included 47712 (54% women) in the longitudinal RHR vs. all-cause new-onset HF analysis, and 47162 participants (55% women) in the RHR trajectory vs. all-cause new-onset HF analysis.

**Figure 1.**
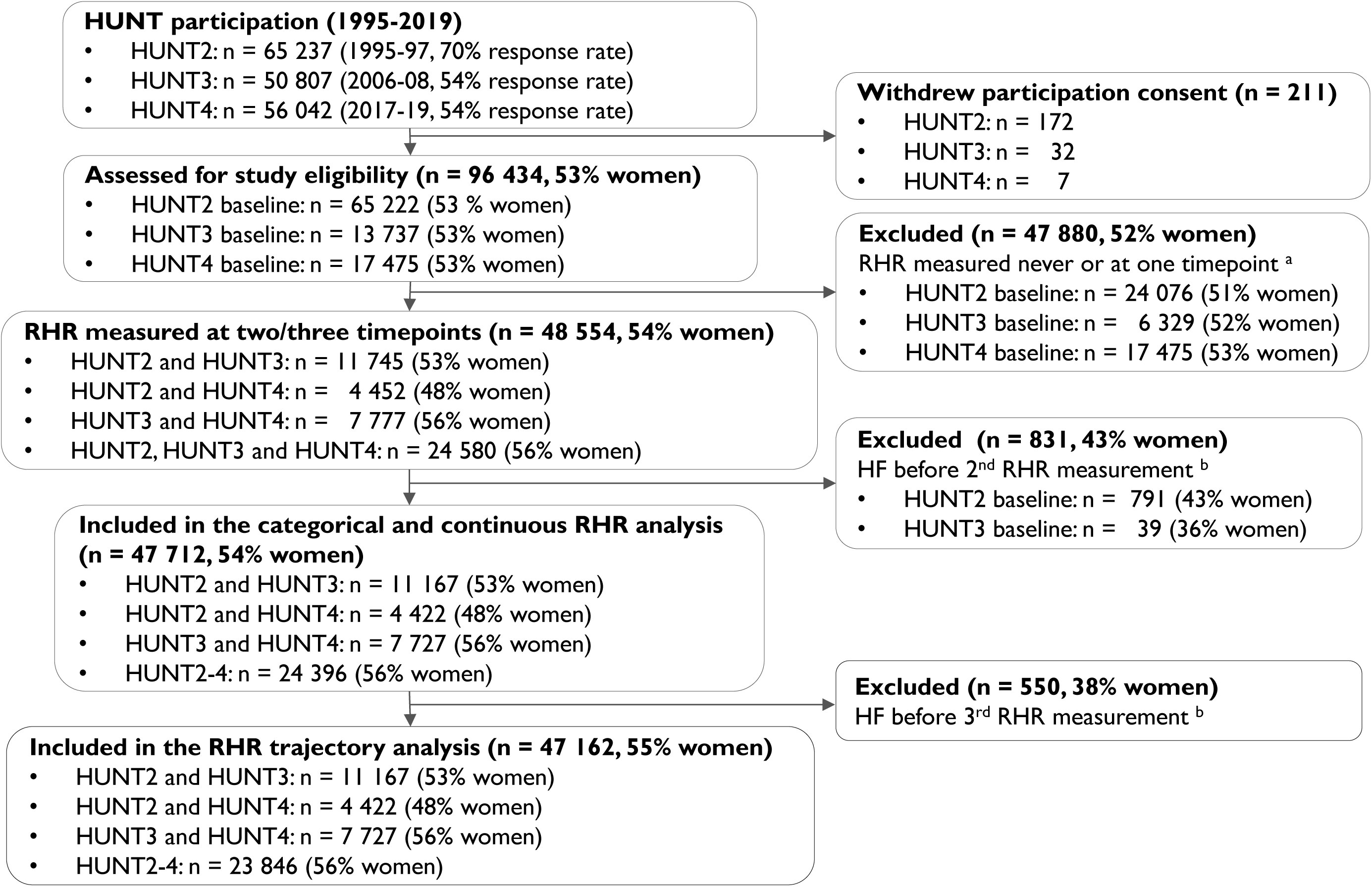
Flow of the participants included in the study. HUNT = The Trøndelag Health Study, RHR = resting heart rate, HF = heart failure, ^a^ RHR set as missing if the participant was pregnant during clinical measurement, ^b^ Incidence HF from hospital journals, the Norwegian Cause of Death Registry, or self-reported HF at baseline HUNT3.

This study followed the Declaration of Helsinki and was approved by the Regional Committee for Medical Research Ethics Central (2018/2416). All participants in HUNT provided their informed written consent.

### Outcome

All-cause incidences of HF during follow-up were identified by data from the Nord-Trøndelag Hospital Trust (i.e. two local hospitals, St. Olavs University hospital) and the Norwegian Cause of Death Registry, using the International Classification of Diseases (ICD)-10 (ICD-9) code I50 (428). HF, hypertensive heart disease and cardiomyopathy diagnoses in hospital journals from 369 participants were previously validated ^21^ according to the current European Society of Cardiology guidelines^22^.

### Clinical investigations

Clinical measurements and venous blood sampling in the HUNT investigations were conducted by trained health professionals^19,20^. Biological material were stored at the HUNT Biobank^23^. RHR and blood pressure were measured three times at 1-min intervals after 2-min seated rest, and before blood sampling, using an arm cuff and the automatic oscillometric method (Dinamap 845XT analyzer, Citikon, Tampa, FL)^19,20,24^. RHR measured during pregnancy was set as missing. We used the mean of the second and third RHR and blood pressure measurements from each HUNT survey in the analysis.

Non-fasting serum cholesterol was measured using the Hitachi 911 Autoanalyzer (Hitachi, Mito, Japan). Hypertension was defined as systolic blood pressure ≥ 140 mmHg and/or diastolic blood pressure ≥ 90 mmHg and/or self-reported use of medication that influences heart rate or blood pressure, and diabetes mellitus was defined as fasting serum glucose ≥ 7.0 mmol/L (HUNT2-3), 2-hour post-load serum glucose ≥ 11.1 mmol/L (HUNT4) or self-reported diabetes mellitus.

### Other covariates

All-cause incidences of atrial fibrillation were identified by hospital data from the Nord-Trøndelag Health Trust and the Norwegian Cause of Death Registry. The HUNT questionnaires contained self-reported data on leisure time physical activity and educational level, smoking, cardiovascular diseases (i.e. stroke, angina pectoris, myocardial infarction) and use of medication for cardiovascular diseases, as well as treatment of hypertension and diabetes mellitus^20^. Educational level was categorized into three: low (Primary school, 7-10 years), middle (1-2 years high school, University qualifying examination) and high (University or other post-secondary education). Participants were categorized as current smokers if they reported smoking daily or occasionally. Physical activity level was categorized into inactive, low, moderate, and high based on a previously published physical activity index from HUNT^25^. We estimated cardiorespiratory fitness as peak oxygen uptake through a HUNT study specific non-exercise regression prediction model based on age, physical activity level, RHR and waist circumference^25^.

### Statistical analysis

Baseline characteristics and variable distribution are presented as absolute numbers and percentages, means ± standard deviations for normally distributed continuous data, and as medians and interquartile range (IQR) if skewed. Multiple imputation by chained equations was used to deal with missing data, with twenty imputed datasets and stratified by sex. The data were assumed to be missing at random. We verified the stability of inference from the imputed datasets by using different missing-data sets (i.e., investigated each imputation step)^26^. All analyses were performed using Stata for Windows, version 18.0, StataCorp LLC, 2023.

In the categorical and continuous analyses, change in RHR was calculated as the difference in mean RHR between the first and second RHR measurement in HUNT2-4. The follow-up period was between the second RHR measurement and the first all-cause new-onset HF, time of death for other causes than HF, or end of follow-up (30 March 2023), whichever came first. We assessed the association between new-onset HF for each 10-bpm change in RHR. Additionally, we categorized changes in RHR according to the mean baseline RHR standard deviation (12 bpm for both sexes) to define categories of decreased, maintained, and increased RHR between the first and second RHR measurement. Restricted cubic splines with five knots were used to display the association between RHR as a continuous variable and new-onset HF.

In the RHR trajectory analysis, we used latent class models to determine groups of RHR trajectories based on up to three RHR measures, using the year of each HUNT-survey as a timescale. We used the Bayesian Information Criterion (BIC) to select the best-fitting model and required that each RHR trajectory included at least 5% of the participants^27^. The model that provided the best fit and had the lowest BIC consisted of three sex-specific trajectory groups with up to a quadratic polynomial type. Models with four or more trajectories led to too small groups (i.e., < 2.5% for women, < 2.7% for men). Posterior predicted probability was used to calculate the probability of each participant belonging to a specific RHR trajectory group. Participants were assigned to one of the three trajectory groups based on the highest probability. For participants who attended all three HUNT surveys, the follow-up period for new-onset HF started after the third RHR measurement timepoint used in the RHR trajectory analysis.

We used Cox proportional hazard models, stratified by sex, and attained age as the time variable, to estimate HR and the 95% CI for the association between changes in RHR and HF, and Schoenfeld residuals to assess the proportional hazard assumption. Cox models were performed on each imputed data set and combined using Rubin’s rule^28^. Variables included in the analyses were chosen a priori based on factors that may affect both RHR and the risk of developing all-cause HF. Model 1 was adjusted for age, Model 2 was adjusted for age, physical activity, body mass index, smoking and baseline RHR, Model 3 was adjusted for Model 2, and hypertension (systolic blood pressure ≥ 140 mmHg, diastolic blood pressure ≥ 90 mmHg or use of antihypertensive medication), diabetes (non-fasting serum glucose ≥ 11.1 mmol/L, HbA1c ≥ 11.1 mmol/L, or self-reported), educational level and total cholesterol, and finally, Model 4 was adjusted for Model 3, atrial fibrillation, self-reported history of stroke, angina pectoris, myocardial infarction and use of medication that may influence heart rate. We used time-updated values in all models. RHR trajectories were not adjusted for baseline RHR. We performed sensitivity analysis for a subsample of participants with validated HF diagnosis from medical record files.

## Results

### Baseline characteristics

Table 1 presents baseline characteristics according to sex and RHR categories. Most of the women and men (74%) maintained their RHR as defined as being within one SD from the first RHR measurement (±12 bpm). Women had 4 bpm higher RHR, and 0.8-1.4 kg/m^2^ lower BMI than men. Furthermore, a higher percentage of women were physically inactive compared to men, while a lower percentage of women had any cardiovascular disease or hypertension. Women and men with a decreased RHR from baseline to the second RHR measurement had a 13-19 bpm higher mean baseline RHR and lower estimated VO_2peak_ (1.4-4.2 mL·min^−1^), compared to participants with a maintained or increased RHR.

**Table 1.** Baseline participant characteristics.

| | Total | Decreased RHR<br>($< -12$ bpm) | Maintained RHR<br>($\pm 12$ bpm) | Increased RHR<br>($> 12$ bpm) |
| --- | --- | --- | --- | --- |
| **Women, n (%)** | 25993 (54%) | 4420 (17%) | 19235 (74%) | 2338 (9%) |
| Age, years (IQR) | 44.6 ± 13.6 | 44.8 ± 14.1 | 44.5 ± 13.4 | 45.3 ± 14.3 |
| Resting heart rate, bpm | 73.8 ± 11.6 | 85.3 (11.5) | 72.0 (10.0) | 67.3 (10.1) |
| SBP, mmHg (IQR) | 125.0 (24.0) | 130.0 (28.0) | 124.0 (23.0) | 125.0 (24.0) |
| DBP, mmHg | 75.9 ± 11.5 | 79.2 ± 12.3) | 75.3 ± 11.2) | 75.3 ± 11.7) |
| Body Mass Index, kg/m<sup>2</sup> (IQR) | 25.2 (5.4) | 25.2 (6.0) | 25.1 (5.3) | 25.4 (5.4) |
| Total cholesterol, mmol/L (IQR) | 5.68 ± 1.26 | 5.76 ± 1.30 | 5.66 ± 1.25 | 5.69 ± 1.23 |
| Smoking, n (%) | 7460 (29%) | 1406 (32%) | 5309 (28%) | 745 (32%) |
| CVD <sup>a</sup>, n (%) | 565 (2%) | 89 (2%) | 410 (2%) | 67 (3%) |
| Hypertension <sup>b</sup>, n (%) | 7209 (28%) | 1664 (38%) | 4871 (25%) | 674 (29%) |
| Diabetes <sup>c</sup>, n (%) | 557 (2%) | 123 (3%) | 367 (2%) | 67 (3%) |
| Physical activity level |  |  |  |  |
| Inactive, n (%) | 4186 (18%) | 660 (16%) | 3115 (18%) | 411 (20%) |
| Low, n (%) | 5993 (25%) | 1030 (26%) | 4445 (25%) | 518 (25%) |
| Moderate, (%) | 6044 (26%) | 1110 (28%) | 4430 (25%) | 504 (25%) |
| High, (%) | 7396 (31%) | 1206 (30%) | 5574 (32%) | 616 (30%) |
| Estimated VO<sub>2peak</sub> <sup>d</sup>, mL·min<sup>-1</sup> (IQR) | 29.2 ± 6.0 | 28.1 ± 6.0 | 29.5 ± 5.9 | 29.4 ± 6.6 |
| Education |  |  |  |  |
| Low (7-10 years primary school), n (%) | 6042 (24%) | 1121 (26%) | 4314 (23%) | 607 (26%) |
| Middle (1-2 years high school), n (%) | 11039 (43%) | 1863 (43%) | 8202 (43%) | 974 (42%) |
| High (University), n (%) | 8566 (33%) | 1353 (31%) | 6500 (34%) | 713 (31%) |
| **Men, n (%)** | 21719 (46%) | 3195 (14%) | 16077 (74%) | 2447 (11%) |
| Age, years (IQR) | 44.9 ± 13.2 | 46.1 ± 13.5 | 44.7 ± 13.0 | 44.7 ± 13.9 |
| Resting heart rate, bpm | 69.6 ± 11.8 | 81.8 (11.5) | 68.2 (10.4) | 63.3 (10.3) |
| SBP, mmHg (IQR) | 134.0 (19.0) | 138.0 (22.0) | 133.0 (20.0) | 133.0 (19.0) |
| DBP, mmHg | 79.8 ± 11.4 | 83.2 ± 12.2 | 79.4 ± 11.1 | 78.3 ± 11.4 |
| Body Mass Index, kg/m<sup>2</sup> (IQR) | 26.2 (4.2) | 26.6 (4.5) | 26.1 (4.1) | 26.2 (4.4) |
| Total cholesterol, mmol/L (IQR) | 5.70 ± 1.13 | 5.88 ± 1.19 | 5.67 ± 1.12 | 5.61 ± 1.14 |
| Smoking, n (%) | 5544 (26%) | 964 (30%) | 3917 (25%) | 663 (27%) |
| CVD <sup>a</sup>, n (%) | 1007 (5%) | 156 (5%) | 701 (4%) | 150 (6%) |
| Hypertension <sup>b</sup>, n (%) | 8722 (40%) | 1648 (52%) | 6130 (38%) | 944 (39%) |
| Diabetes <sup>c</sup>, n (%) | 565 (3%) | 114 (4%) | 373 (2%) | 78 (3%) |
| Physical activity level |  |  |  |  |
| Inactive, n (%) | 2526 (13%) | 419 (14%) | 1827 (13%) | 280 (13%) |
| Low, n (%) | 3907 (20%) | 602 (21%) | 2896 (20%) | 409 (19%) |
| Moderate, (%) | 4959 (25%) | 767 (27%) | 3673 (25%) | 519 (24%) |
| High, (%) | 8224 (42%) | 1105 (38%) | 6191 (42%) | 928 (43%) |
| Estimated VO<sub>2peak</sub> <sup>d</sup>, mL·min<sup>-1</sup> (IQR) | 38.1 ± 6.8 | 35.6 ± 6.7 | 38.4 ± 6.7 | 39.1 ± 7.3 |
| Education |  |  |  |  |
| Low (7-10 years primary school), n (%) | 3761 (17%) | 644 (20%) | 2702 (17%) | 415 (17%) |
| Middle (1-2 years high school), n (%) | 11291 (52%) | 1634 (52%) | 8369 (52%) | 1288 (53%) |
| High (University), n (%) | 6473 (30%) | 883 (28%) | 4876 (31%) | 714 (30%) |

Baseline characteristics by change in resting heart rate (beats per minute (bpm)) from baseline to the 2<sup>nd</sup> resting heart rate measure. Values are presented as means ( ± SD), No. (percentages) or median (interquartile range (IQR)). <sup>a</sup> CVD: self-reported stroke, angina pectoris, myocardial infarction, use of antiarrhythmic agents. <sup>b</sup> Hypertension: systolic blood pressure (SBP) ≥ 140 mmHg, diastolic blood pressure (DBP) ≥ 90 mmHg or use of medication that influences heart rate or blood pressure. <sup>c</sup> Diabetes: non-fasting serum glucose ≥ 11.1 mmol/L, HbA1c ≥ 11.1 mmol/L or self-reported diabetes in questionnaire. <sup>d</sup> VO<sub>2peak</sub>: estimated peak oxygen
consumption.

Women and men in the high RHR trajectory had a 2.7-6.1 mL·min^−1^ lower estimated baseline VO_2peak_ compared to those in the low or moderate RHR trajectories (Supplementary data, Table S1). Furthermore, a higher proportion of participants had hypertension and were current smokers in the high RHR trajectory group compared to the lowest or moderate trajectory groups. Women were more likely than men to have a high baseline RHR, where 9% of women and 5% of men had RHR ≥ 85 bpm. Moreover, a higher proportion of women and men with a high baseline RHR were current smokers, hypertensive, and inactive compared to the proportion of participants with a low or normal baseline RHR (Supplementary data, Table S2).

### Longitudinal characteristics

The average observation period for RHR was 12.0 ± 3.2 years between baseline (HUNT2 or HUNT3) and the second RHR measurement (HUNT3 or HUNT4), and 10.6 ± 0.7 years between the second and third RHR measurements (HUNT3 to HUNT4). Of the 47713 participants with a total of 586542 person-years of follow-up (mean 12.3 years, maximum 16.5 years), 2880 were diagnosed with all-cause HF (48% women, Table 2). Women with all-cause HF were diagnosed at an older age (median (IQR), 82 (74-88) years) than men (78 (69-84) years) and had a lower overall HF incidence rate (Table 2), with an incidence rate ratio of 0.67 (95% CI: 0.57-0.77).

**Table 2.**
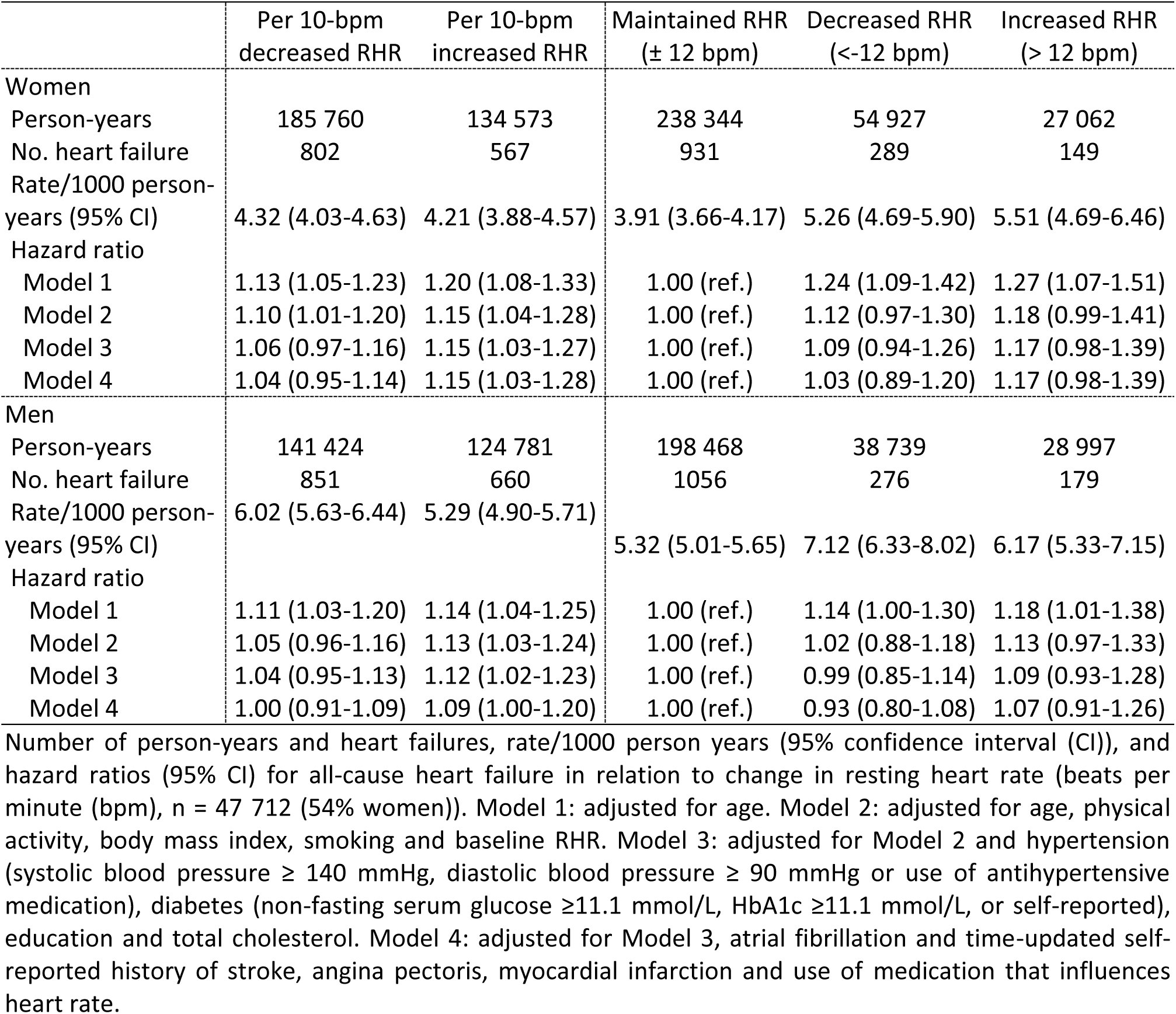
Change in resting heart rate and heart failure risk.

Among the 47162 participants with a total of 328652 person-years (mean 7.0 ± 4.1 years, maximum 16.5 years) of follow-up included in the RHR trajectory analysis, 2330 participants (50% women) were diagnosed with all-cause HF (Table 3). Women’s RHR trajectories were consistently 3.7-4.8 bpm higher than men’s (Figure 2). There was a nearly equal distribution of women and men in the low and moderate RHR trajectories, while 5% of women and 8% of men belonged to the high RHR trajectory (Table 3).

**Figure 2.**
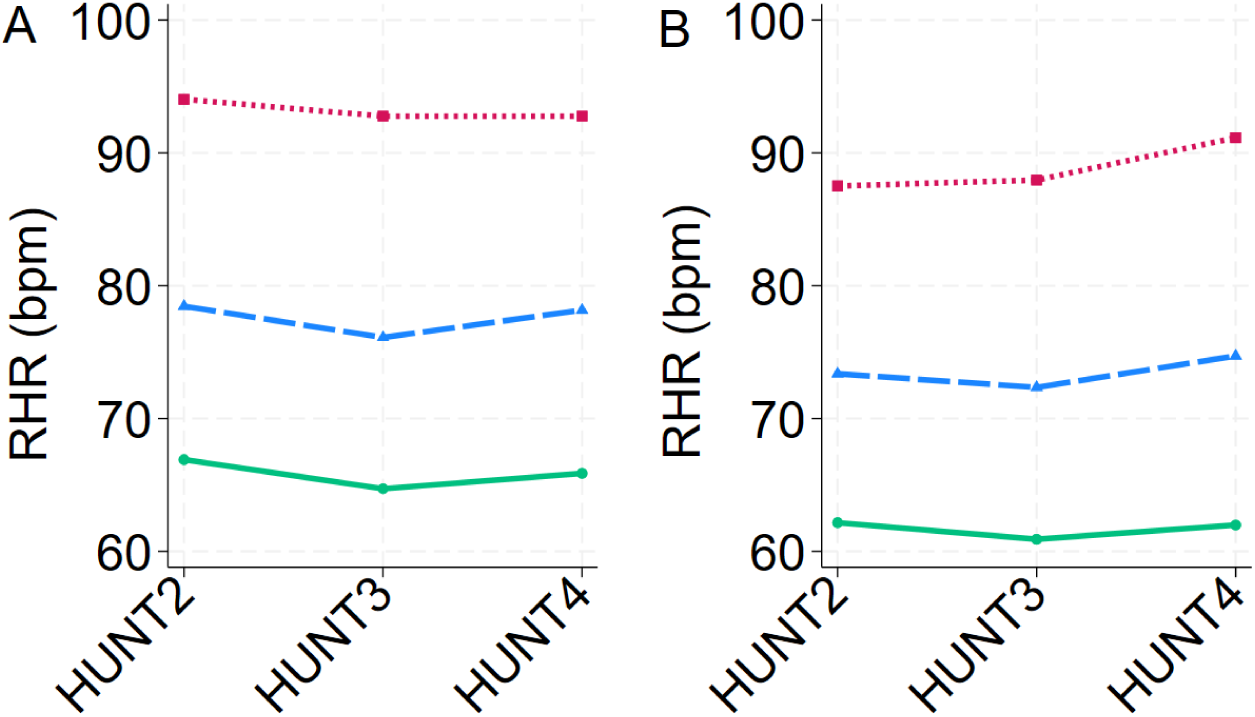
Resting heart rate trajectories in women (A) and men (B), The Trøndelag Health Study, 1995-2019. Sex-specific means of resting heart rate (beats per minute) presented according to three HUNT surveys and three resting heart rate trajectory groups. Latent class models determined resting heart rate trajectory groups (Stata 18 traj).

**Table 3.** Resting heart rate trajectories and heart failure risk.

|  | Resting heart rate trajectories |  |  |
| --- | --- | --- | --- |
|  | Low | Moderate | High |
| Women | 63.9-66.1 bpm | 77.0-79.3 bpm | 94.0-96.0 bpm |
| No. participants | 12 353 (48%) | 12 099 (47%) | 1 333 (5%) |
| Person-years | 84122 | 83150 | 9351 |
| No. heart failures | 525 | 544 | 92 |
| Rate/1000 person-years (95% CI) | 6.24 (5.73-6.80) | 6.54 (6.02-7.12) | 9.84 (8.02-12.07) |
| Hazard ratio |  |  |  |
| Model 1 | 1.00 (ref.) | 1.01 (0.90-1.14) | 1.48 (1.19-1.85) |
| Model 2 | 1.00 (ref.) | 0.96 (0.85-1.09) | 1.34 (1.07-1.68) |
| Model 3 | 1.00 (ref.) | 0.96 (0.85-1.09) | 1.32 (1.06-1.65) |
| Model 4 | 1.00 (ref.) | 1.00 (0.89-1.13) | 1.43 (1.14-1.79) |
| Men | 59.9-61.3 bpm | 73.3-75.4 bpm | 89.2-92.6 bpm |
| No. participants | 9 511 (44%) | 10 163 (48%) | 1 704 (8%) |
| Person-years | 65772 | 73747 | 12510 |
| No. heart failures | 498 | 539 | 132 |
| Rate/1000 person-years (95% CI) | 7.57 (6.93-8.27) | 7.31 (6.72-7.95) | 10.55 (8.90-12.51) |
| Hazard ratio |  |  |  |
| Model 1 | 1.00 (ref.) | 1.14 (1.01-1.29) | 1.56 (1.28-1.89) |
| Model 2 | 1.00 (ref.) | 1.07 (0.95-1.22) | 1.42 (1.17-1.73) |
| Model 3 | 1.00 (ref.) | 1.06 (0.93-1.20) | 1.36 (1.12-1.66) |
| Model 4 | 1.00 (ref.) | 1.08 (0.96-1.23) | 1.41 (1.16-1.72) |

Number of person-years and heart failures, rate/1000 person years (95% confidence interval (CI)), and hazard ratios (95% CI) for all-cause heart failure in relation to trajectories groups of resting heart rate (beats per minute (bpm), n = 47 162 (55% women)). Model 1: adjusted for age. Model 2: adjusted for age, physical activity, body mass index and smoking. Model 3: adjusted for Model 2 and hypertension (systolic blood pressure ≥ 140 mmHg, diastolic blood pressure ≥ 90 mmHg or use of antihypertensive medication), diabetes (non-fasting serum glucose ≥ 11.1 mmol/L, HbA1c ≥ 11.1 mmol/L, or self-reported), education and total cholesterol. Model 4: adjusted for Model 3, atrial fibrillation and time-updated self-reported history of stroke, angina pectoris, myocardial infarction and use of medication that influences heart rate.

Nelson-Aalen cumulative hazard estimates indicated a higher hazard of HF in participants older than 80 years compared to younger participants, and in men compared to women (Supplementary data, Figure S1). From baseline to the second RHR measure, the overall mean change in RHR was -2 ± 12 bpm for women, and -1 ± 12 bpm for men (Supplementary data, Figure S2). The maximum decrease in RHR from baseline to the second RHR measure was -68 bpm for women and -67 for men, and the maximum increase was 74 bpm for both sexes (Supplementary data, Figure S2). Nineteen participants (26% women) had an increase in RHR of 50 bpm or more.

### Categorical and continuous change in RHR and HF risk

In fully adjusted categorical analysis, women with an increase in RHR had a trend of 17% higher risk of new-onset HF compared to women with a stable RHR (Table 2). No such trend was found for men. In continuous adjusted linear analyses of change in RHR, each 10-bpm increase in RHR was associated with an 15% higher risk of HF in women and a 9% higher risk in men (Table 2). There was no association between a decrease in RHR for either sex, neither in the categorical nor in the continuous analysis. Excluding 59 participants who were re-categorized from HF to non-valid HF in HUNT4HOPE did not affect the risk estimates (Supplementary data, Table S3).

The adjusted restricted cubic splines displayed a non-linear relationship between RHR and HF, indicating a higher risk of HF above certain increase in RHR (e.g. above 25 bpm for women and 32 bpm for men (Figure 3). Although not significant for a decrease in RHR, the dose-response relationship appeared J-shaped for women, without a prominent J-shaped curve for men (Figure 3). Both sexes had wide confidence intervals for large changes in RHR (i.e. above 30 bpm change).

**Figure 3.**
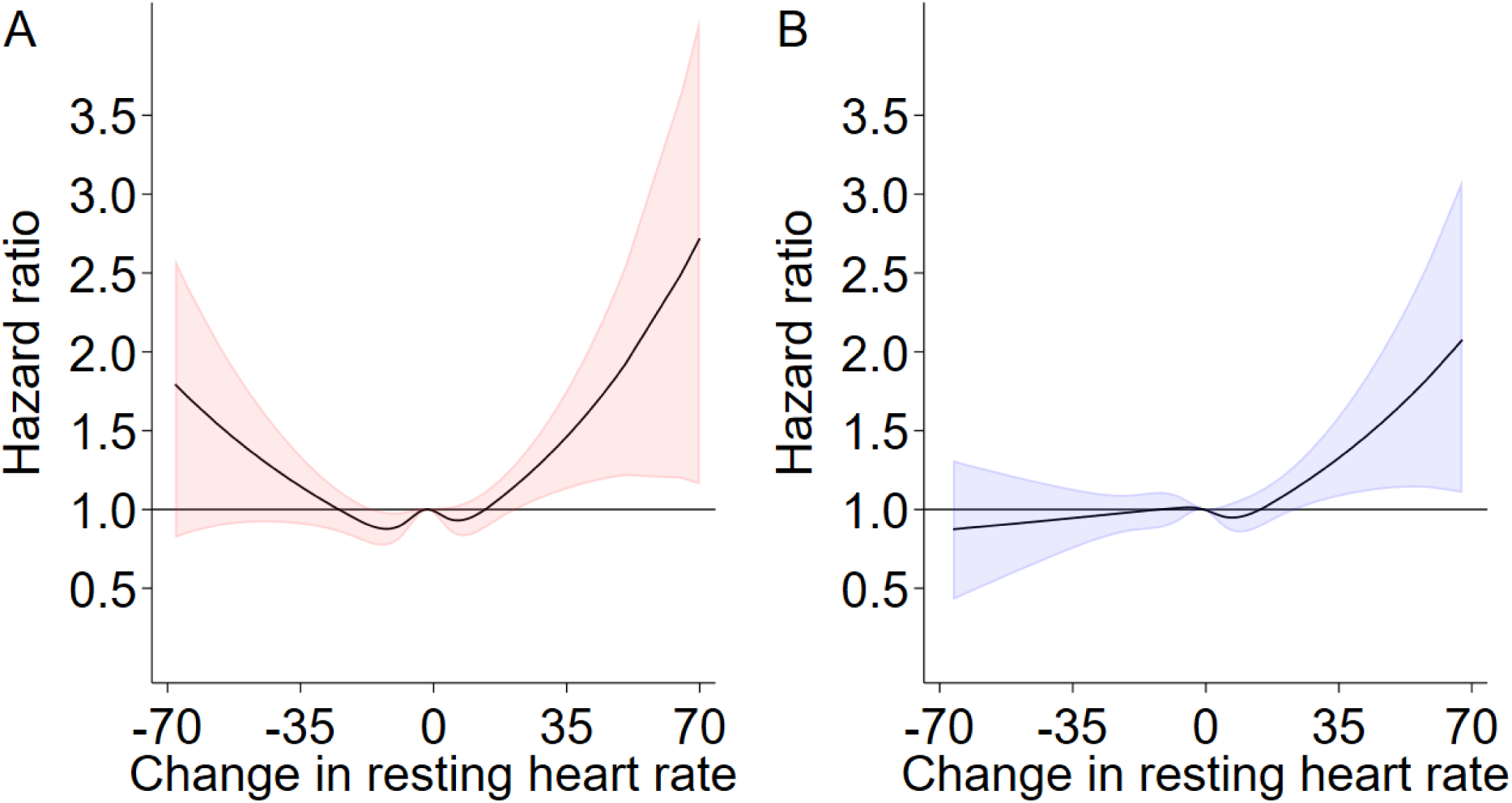
Restricted cubic splines displaying the association between resting heart rate and the risk of incidence of all-cause heart failure for women (A) and men (B). Sex-specific hazard ratios and 95% confidence intervals presented according to change in resting heart rate (beats per minute) from baseline to follow-up. Restricted cubic splines with five knots were used to model the non-linear relationship between hazard ratio and change in resting heart rate (Stata 18 mkspline). Adjusted for age, physical activity, body mass index, smoking, hypertension (systolic blood pressure ≥ 140 mmHg, diastolic blood pressure ≥ 90 mmHg or use of antihypertensive medication), diabetes (non-fasting serum glucose ≥ 11.1 mmol/L, HbA1c ≥ 11.1 mmol/L, or self-reported), education, total cholesterol, atrial fibrillation and time-updated self-reported history of stroke, angina pectoris, myocardial infarction and use of medication that influence heart rate or blood pressure.

Adjusting for lifestyle and known HF risk factors (e.g. hypertension, total cholesterol and obesity) gave 5-7% reduction in risk estimates for women and 1-4% reduction for men per 10-bpm increase in RHR (Table 2). Further adjustments for cardiovascular diseases had a similar effect on the hazard ratios for both women and men, despite women showing a slightly higher reduction in models 2 and 3 (Table 2).

### RHR trajectories and HF risk

In fully adjusted models, women and men in the high RHR trajectories had 43% and 41% higher risk of new-onset HF, compared with the lowest RHR trajectory (Table 3). A visual summary of the results is presented in the graphical abstract. There was no association between a moderate RHR trajectory and HF when compared to the low RHR trajectory. Adjusting for lifestyle factors had a similar impact on the hazard ratio for women and men with a high RHR trajectory (i.e. 16-20% risk reduction), while adjustment for known risk factors for HF and cardiovascular diseases had a larger impact on the hazard ratios for women with a high RHR trajectory compared to men (Table 3). Specifically, the risk for women increased from 32% in lifestyle-adjusted model to 43% in fully adjusted model, while for men it increased from 36% to 41% (Table 3).

### Baseline HF risk

Women and men with a high baseline RHR (≥ 85 bpm) had 19% and 23% higher risk of new-onset HF, respectively, compared with a normal baseline RHR (60-84 bpm). There was no association between a low baseline RHR (< 60 bpm) and new-onset HF for neither women nor men (Supplementary data, Table S4). In adjusted continuous linear analyses, Schoenfeld residuals indicated violation of the proportional hazard assumption for each 10-bpm increase in RHR if the RHR started at 50 bpm. There was no violation of proportional hazard assumption when starting at 60 bpm. Each 10-bpm higher baseline RHR, starting from 60-bpm, was associated with a trend of 4% higher risk of new-onset HF for women, and a statistically significant 9% higher risk for men after covariate adjustments (Supplementary data, Table S4).

## Discussion

In this study evaluating the association between longitudinal RHR and new-onset HF in a Norwegian cohort of 47713 adults, the main findings indicated a similarly higher risk of HF in both women and men with an increased RHR or high RHR trajectory. Each 10-bpm increase in RHR was associated with a 13% higher risk of HF for women and 9% higher for men. We identified three sex-specific trajectory groups reflecting individual RHR patterns across up to 33 years of aging, with a 43% higher risk for women and a 42% higher for men with a high RHR trajectory compared to a low RHR trajectory. Our findings indicate that a large increase in RHR or a consistently high RHR may be early warning signs of HF for both sexes.

### Comparison with literature

#### Increase in RHR

The finding of a 15% higher risk of HF in women and 10% in men per each 10-bpm increase in RHR aligns with two American studies^16,17^. However, our risk estimates were lower. One study reported a 13% (95% CI: 9-16%) higher risk of HF per 5-bpm increase in RHR^16^, and the other found a 38% (95% CI: 2-86%) higher risk per 10-bpm increase^17^ in combined analyses of both sexes. Differences in absolute risk, exclusion criteria, and methods of data collection may explain these discrepancies^16,17^. The incidence of HF in Norway rate is lower than in the USA (5.04-5.40 vs. 6.0-7.9 per 1000-person-years)^29,30^, which aligns with the 6% of participants who developed HF in our study compared to the 18.7% reported in one American study^16^. The other study^17^ reported a 1.7% incidence of HF but may have introduced selection bias by excluding participants with any cardiovascular event prior to the final RHR measurement. Additionally, it assessed RHR via palpation^17^, a method more prone to measurement error than the automatic oscillometric method^31^ used in HUNT. Despite these differences, our findings support previous findings^16,17^ that increased RHR is associated with a similarly elevated risk of HF in women and men.

#### RHR trajectories

While we observed a 41-43% higher risk of HF for both women and men with a high RHR trajectory, a US study reported higher risk of HF for women (HR=2.73 (95% CI: 1.51-4.95) than for men (HR=2.37 (95% CI: 1.44-3.91) within the same trajectory^18^. This study included 3412 participants (54% women) free of known HF with a mean age of 67 years at baseline and followed the participants for 10 years^18^. Similarly to our study, 8.1% of the participants developed HF during follow-up^18^. Participants in our study may have had a more favorable baseline risk profile^5,8^, given that the participants in the American study were, on average 23 years older, had a mean BMI that was 2.6 kg/m² higher, and 52% of the women were hypertensive^18^, compared to 28% in our study. Despite differences in baseline risk profiles, both studies identified a strong association between RHR trajectories and HF. This underscores the importance of longitudinal RHR as a crucial biomarker for heart health.

#### Decrease in RHR

The absence of an association between a decrease in RHR and new-onset HF both aligns with and contrasts previous studies^16,17^. The lack of statistical significance in the association between decreased RHR and HF aligns with a previous study observing RHR over five years^17^. In a combined analysis using restricted cubic splines for both women and men, a 20% lower risk of incident HF was detected with a decreased RHR^16^. Variations in the covariates included in the analysis may explain the different findings between studies. Our study adjusted for HF risk factors such as total cholesterol, stroke, and angina pectoris^3^, which the other study did not^16^. Additionally, we used time-updated values for physical activity and smoking, rather than baseline values^16^. Using time-updated values may affect risk estimates, as regular physical activity can lower RHR and reduce HF risk^8^. For example, one study found a 91% higher risk of incidence HF for unfit men compared to fit men^32^. Further research is needed to understand potential clinical implications of decreased RHR.

#### Single time-point RHR vs. longitudinal RHR

A small proportion of participants, i.e. 5% of the women and 8% of the men, had increased risk of HF associated with up to thirty years of elevated RHR. We found a stronger association between longitudinal RHR and incident HF compared to baseline RHR data alone, suggesting that repeated RHR measurements might be particularly important in individuals with consistently elevated RHR to enhance HF risk assessment and implement preventive measures. While a single RHR measurement can reflect underlying cardiovascular pathology, repeated measurements can capture long-term patterns and changes in physiological and environmental risk factors for HF ^7^. Additionally, RHR trajectories provide more information by incorporating a third RHR measurement into the analysis, as well as providing longer exposure time. With more data points, a 33-year observation period, and older participants at the start of follow-up, we obtained higher risk estimates for women and men compared to using only two RHR measurements. HF often develops gradually in both sexes due to cumulative exposure to risk factors^3^, and including multiple RHR measurements offers a comprehensive view of individual RHR trends over time.

### Incidence rates and risk factors for HF

In our study, the incidence rate of HF per 1000 person-years was 5.0 for women and 6.8 for men. These rates aligns with findings from a study of German men^33^ but are higher than the overall estimate from 12 European countries, which was 3.2 per 1000 person-years^34^. Potential mechanisms for the lower incidence rate of HF in women compared to men include differences in risk factors such as age, diabetes mellitus, and hypertension^3^. European women has been shown to develop HF 4-6 years later than men^2,35^, a finding confirmed in our study where median age at HF diagnosis was 84 years for women and 80 years for men. Aging is related to physiological changes in heart structure, such as increased left ventricular wall thickness and decreased left ventricular mass, and these changes might be sex-specific^36^. Additionally, a recent European study found that hypertension, total cholesterol, and obesity contributed more to HF risk in women than in men^37^. For example, a study reported that women with diabetes mellitus had a five-fold higher risk of HF compared to a two-fold risk for men, and women with hypertension had a three-fold higher risk compared to a two-fold risk for men^3^. The risk factor adjustments in our models revealed a consistent pattern, showing that the known risk factors for HF such as hypertension, total cholesterol, and obesity had a greater impact for our risk models in women compared to men. Overall, the lower incidence rate of HF in women compared to men in our study may be attributed to both biological differences and diverse risk factor profiles.

### Strengths and limitations

The main strengths of this study include a well-characterized, large cohort of 47713 participants and a rigorous methodology which combines categorical, continuous, and trajectory-based analyses supported by comprehensive data on key heart failure risk factors (e.g., physical activity, blood pressure and cardiovascular diseases). Importantly, this research provides long-term evidence on RHR trajectories and incident HF within a European population, where such studies have been notably scarce.

The equal representation of women and men enabled sex-specific analyses, provided high statistical power, and minimized the risk of type II errors^38^. The HUNT Study facilitates longitudinal comparisons through repeated clinical measurements, with questionnaires covering various topics and consistently maintain items across surveys^19^. Additionally, the intervals between RHR measurements in our study was 7-9 years longer than in previous studies^16,17^, enabling the use of more recent, time-updated values for RHR as well as potential confounders in our analysis. Misclassification is a potential limitation in epidemiological studies^38^. However, our cohort data was linked to national health registers using the national personal identification numbers and high-quality hospital journals for HF diagnosis, significantly reducing the risk of misclassification. HF was confirmed in 310 of the 369 participants evaluated in HUNT4HOPE study. Excluding participants with non-valid HF did not affect our risk estimates. However, we had validation data for only 369 participants, and HF subtype data for 224 participants, out of the 2880 participants who developed HF. Thus, it remains unclear whether the underlying case of elevated RHR is due to physiological stimulation, such as increased sympathetic load, or if it serves as an early warning sign for pathological remodeling, characterized by significant decreased stroke volume and a compensatory increase in RHR to maintain necessary cardiac output.

## Conclusion

All-cause HF was similarly associated with increased RHR and a high RHR trajectory for women and men. Estimating HF risk using RHR trajectories provided a stronger association between RHR and HF compared to using a single RHR measurement.

### Clinical implications

Repeated RHR measurements have the potential to be utilized more extensively as a low-cost clinical tool for monitoring heart disease risk over decades. Knowledge of longitudinal RHR might improve prevention or initiate relevant treatments in high-risk individuals. With the advent of new and more accurate wrist worn devices for measuring heart rate, the potential for home monitoring of high-risk individuals is substantial, potentially reducing the patient load on health services. Repeated RHR measurements might also provide valuable prognostic insights by minimizing the impact of temporary heart rate fluctuations. These measurements can help identify the small proportion of men and women in a population with consistently elevated RHR and a high risk of developing HF. Future studies, as well as clinical investigations should explore whether the underlying cause of elevated RHR is physiological or pathological. Additionally, long term RHR measurements can reveal changes in HF risk profiles that might not be detectable with a single RHR measurement and highlight individuals with a high RHR who should be further evaluated.

## Data Availability

The data that support the findings of this study are not publicly available due to privacy restrictions.

## Acknowledgements

The Trøndelag Health Study (HUNT) is a collaboration between the HUNT Research Centre (Faculty of Medicine and Health Sciences, Norwegian University of Science and Technology (NTNU)), Trøndelag County Council, Central Norway Regional Health Authority, and the Norwegian Institute of Public Health. Thank you to the participants in HUNT. We want to thank clinicians and other employees at Nord-Trøndelag Hospital Trust for their support and for contributing to data collection in this research project.

## Sources of Funding

Sources of support, for all authors, for the work: The study was funded by Nord University.

## Disclosures

Conflict of interest: None.

## Authors’ Contributions

All authors contributed to the conception and/or design of the work. TK and HD contributed to the acquisition of data for the work. TBB and LMSH ran the analysis. All authors contributed to the interpretation of data. LMSH drafted the first version of the manuscript. All authors critically revised the manuscript, gave final approval, and agreed to be accountable for all aspects of work ensuring integrity and accuracy.

**Supplementary data, Figure S1.** Nelson-Aalen cumulative hazard estimates of heart failure. Sex-specific cumulative hazard presented according to age (years) for women (A, solid red line) and men (B, dotted blue line).

**Supplementary data, Figure S2**. Density plot for change in resting heart rate from baseline to follow-up. Sex-specific density presented according to change in resting heart rate (beats per minute) across a median of 12.2 years of aging for women (solid line) and men (dotted line).

## Notes

Sources of support: The study was funded by Nord University, Bodø/Levanger, Norway

### Competing Interest Statement

The authors have declared no competing interest.

### Clinical Trial

Not applicable

### Funding Statement

The study was funded by Nord University, a government-funded institution.

## References

1. Disease GBD, Injury I, Prevalence C. Global, regional, and national incidence, prevalence, and years lived with disability for 354 diseases and injuries for 195 countries and territories, 1990-2017: a systematic analysis for the Global Burden of Disease Study 2017. Lancet. 2018;392:1789–1858. doi: 10.1016/S0140-6736(18)32279-7

2. Bleumink GS, Knetsch AM, Sturkenboom MC, Straus SM, Hofman A, Deckers JW, Witteman JC, Stricker BH. Quantifying the heart failure epidemic: prevalence, incidence rate, lifetime risk and prognosis of heart failure The Rotterdam Study. Eur Heart J. 2004;25:1614–1619. doi: 10.1016/j.ehj.2004.06.038

3. Lala A, Tayal U, Hamo CE, Youmans Q, Al-Khatib SM, Bozkurt B, Davis MB, Januzzi J, Mentz R, Sauer A, et al. Sex Differences in Heart Failure. J Card Fail. 2022;28:477–498. doi: 10.1016/j.cardfail.2021.10.006

4. Regitz-Zagrosek V. Sex and Gender Differences in Heart Failure. Int J Heart Fail. 2020;2:157–181. doi: 10.36628/ijhf.2020.0004

5. McDonagh TA, Metra M, Adamo M, Gardner RS, Baumbach A, Bohm M, Burri H, Butler J, Celutkiene J, Chioncel O, et al. 2023 Focused Update of the 2021 ESC Guidelines for the diagnosis and treatment of acute and chronic heart failure. Eur Heart J. 2023;44:3627–3639. doi: 10.1093/eurheartj/ehad195

6. Aune D, Sen A, ó’Hartaigh B, Janszky I, Romundstad PR, Tonstad S, Vatten LJ. Resting heart rate and the risk of cardiovascular disease, total cancer, and all-cause mortality – A systematic review and dose–response meta-analysis of prospective studies. *Nutrition*, Metabolism and Cardiovascular Diseases. 2017;27:504–517. doi: 10.1016/j.numecd.2017.04.004

7. Olshansky B, Ricci F, Fedorowski A. Importance of resting heart rate. Trends Cardiovasc Med. 2022. doi: 10.1016/j.tcm.2022.05.006

8. Valentini M, Parati G. Variables influencing heart rate. Prog Cardiovasc Dis. 2009;52:11–19. doi: 10.1016/j.pcad.2009.05.004

9. Shi Y, Zhou W, Liu X, Ping Z, Li YQ, Wang C, Lu J, Mao ZX, Zhao J, Yin L, et al. Resting heart rate and the risk of hypertension and heart failure: a dose-response meta-analysis of prospective studies. J Hypertens. 2018;36:995–1004. doi: 10.1097/hjh.0000000000001627

10. Nauman J, Nilsen TI, Wisloff U, Vatten LJ. Combined effect of resting heart rate and physical activity on ischaemic heart disease: mortality follow-up in a population study (the HUNT study, Norway). J Epidemiol Community Health. 2010;64:175–181. doi: 10.1136/jech.2009.093088

11. Seviiri M, Lynch BM, Hodge AM, Yang Y, Liew D, English DR, Giles GG, Milne RL, Dugue PA. Resting heart rate, temporal changes in resting heart rate, and overall and cause-specific mortality. Heart. 2018;104:1076–1085. doi: 10.1136/heartjnl-2017-312251

12. Sharashova E, Wilsgaard T, Løchen ML, Mathiesen EB, Njølstad I, Brenn T. Resting heart rate trajectories and myocardial infarction, atrial fibrillation, ischaemic stroke and death in the general population: The Tromsø Study. Eur J Prev Cardiol. 2017;24:748–759. doi: 10.1177/2047487316688983

13. Vazir A, Claggett B, Jhund P, Castagno D, Skali H, Yusuf S, Swedberg K, Granger CB, McMurray JJ, Pfeffer MA, et al. Prognostic importance of temporal changes in resting heart rate in heart failure patients: an analysis of the CHARM program. Eur Heart J. 2015;36:669–675. doi: 10.1093/eurheartj/ehu401

14. Vazir A, Claggett B, Pitt B, Anand I, Sweitzer N, Fang J, Fleg J, Rouleau J, Shah S, Pfeffer MA, et al. Prognostic Importance of Temporal Changes in Resting Heart Rate in Heart Failure and Preserved Ejection Fraction: From the TOPCAT Study. JACC Heart Fail. 2017;5:782–791. doi: 10.1016/j.jchf.2017.08.018

15. Wei CC, Shyu KG, Chien KL. Association of heart rate trajectory patterns with the risk of adverse outcomes for acute heart failure in a heart failure cohort in Taiwan. Acta Cardiologica Sinica. 2020;36:439–447. doi: 10.6515/ACS.202009_36(5).20200519A

16. Vazir A, Claggett B, Cheng S, Skali H, Shah A, Agulair D, Ballantyne CM, Vardeny O, Solomon SD. Association of Resting Heart Rate and Temporal Changes in Heart Rate With Outcomes in Participants of the Atherosclerosis Risk in Communities Study. JAMA Cardiol. 2018;3:200–206. doi: 10.1001/jamacardio.2017.4974

17. Nwabuo CC, Appiah D, Moreira HT, Vasconcellos HD, Aghaji QN, Ambale-Venkatesh B, Rana JS, Allen NB, Lloyd-Jones DM, Schreiner PJ, et al. Temporal Changes in Resting Heart Rate, Left Ventricular Dysfunction, Heart Failure and Cardiovascular Disease: CARDIA Study. Am J Med. 2020;133:946–953. doi: 10.1016/j.amjmed.2019.12.035

18. Guardino CE, Pan S, Vasan RS, Xanthakis V. Multi-system trajectories and the incidence of heart failure in the Framingham Offspring Study. PLoS One. 2022;17:e0268576. doi: 10.1371/journal.pone.0268576

19. Asvold BO, Langhammer A, Rehn TA, Kjelvik G, Grontvedt TV, Sorgjerd EP, Fenstad JS, Heggland J, Holmen O, Stuifbergen MC, et al. Cohort Profile Update: The HUNT Study, Norway. International Journal of Epidemiology. 2022. doi: 10.1093/ije/dyac095

20. Krokstad S, Langhammer A, Hveem K, Holmen TL, Midthjell K, Stene TR, Bratberg G, Heggland J, Holmen J. Cohort Profile: the HUNT Study, Norway. Int J Epidemiol. 2013;42:968–977. doi: 10.1093/ije/dys095

21. Rye CS, Ofstad AP, Asvold BO, Romundstad PR, Horn J, Dalen H. The influence of diagnostic subgroups, patient- and hospital characteristics for the validity of cardiovascular diagnoses-Data from a Norwegian hospital trust. PLoS One. 2024;19:e0302181. doi: 10.1371/journal.pone.0302181

22. McDonagh TA, Metra M, Adamo M, Gardner RS, Baumbach A, Bohm M, Burri H, Butler J, Celutkien J, Chioncel O, et al. 2021 ESC Guidelines for the diagnosis and treatment of acute and chronic heart failure Developed by the Task Force for the diagnosis and treatment of acute and chronic heart failure of the European Society of Cardiology (ESC) With the special contribution of the Heart Failure Association (HFA) of the ESC. Eur J Heart Fail. 2022;24:4–131. doi: 10.1002/ejhf.2333

23. Næss M, Kvaloy K, Sorgjerd EP, Saetermo KS, Noroy L, Rostad AH, Hammer N, Alto TG, Vikdal AJ, Hveem K. Data Resource Profile: The HUNT Biobank. Int J Epidemiol. 2024;53. doi: 10.1093/ije/dyae073

24. Lund-Larsen PG. Blood pressure measured with a sphygmomanometer and with Dinamap under field conditions – a comparison. 1997;7:235–241. doi: 10.5324/nje.v7i2.413

25. Nes BM, Janszky I, Vatten LJ, Nilsen TI, Aspenes ST, Wisløff U. Estimating V·O 2peak from a nonexercise prediction model: the HUNT Study, Norway. Med Sci Sports Exerc. 2011;43:2024–2030. doi: 10.1249/MSS.0b013e31821d3f6f

26. White IR, Royston P, Wood AM. Multiple imputation using chained equations: Issues and guidance for practice. Stat Med. 2011;30:377–399. doi: 10.1002/sim.4067

27. Herle M, Micali N, Abdulkadir M, Loos R, Bryant-Waugh R, Hubel C, Bulik CM, De Stavola BL. Identifying typical trajectories in longitudinal data: modelling strategies and interpretations. Eur J Epidemiol. 2020;35:205–222. doi: 10.1007/s10654-020-00615-6

28. White IR, Royston P. Imputing missing covariate values for the Cox model. Stat Med. 2009;28:1982–1998. doi: 10.1002/sim.3618

29. Savarese G, Becher PM, Lund LH, Seferovic P, Rosano GMC, Coats AJS. Global burden of heart failure: a comprehensive and updated review of epidemiology. Cardiovasc Res. 2023;118:3272. doi: 10.1093/cvr/cvac013

30. Odegaard KM, Lirhus SS, Melberg HO, Hallen J, Halvorsen S. A nationwide registry study on heart failure in Norway from 2008 to 2018: variations in lookback period affect incidence estimates. BMC Cardiovasc Disord. 2022;22:88. doi: 10.1186/s12872-022-02522-y

31. Gudmundsdottir KK, Fredriksson T, Svennberg E, Al-Khalili F, Friberg L, Habel H, Frykman V, Engdahl J. Performance of pulse palpation compared to one-lead ECG in atrial fibrillation screening. Clin Cardiol. 2021;44:692–698. doi: 10.1002/clc.23595

32. Myers J, Kokkinos P, Chan K, Dandekar E, Yilmaz B, Nagare A, Faselis C, Soofi M. Cardiorespiratory Fitness and Reclassification of Risk for Incidence of Heart Failure: The Veterans Exercise Testing Study. Circ Heart Fail. 2017;10. doi: 10.1161/circheartfailure.116.003780

33. Störk S, Handrock R, Jacob J, Walker J, Calado F, Lahoz R, Hupfer S, Klebs S. Epidemiology of heart failure in Germany: a retrospective database study. Clin Res Cardiol. 2017;106:913–922. doi: 10.1007/s00392-017-1137-7

34. Seferovic PM, Vardas P, Jankowska EA, Maggioni AP, Timmis A, Milinkovic I, Polovina M, Gale CP, Lund LH, Lopatin Y, et al. The Heart Failure Association Atlas: Heart Failure Epidemiology and Management Statistics 2019. Eur J Heart Fail. 2021;23:906–914. doi: 10.1002/ejhf.2143

35. Odegaard KM, Hallen J, Lirhus SS, Melberg HO, Halvorsen S. Incidence, prevalence, and mortality of heart failure: a nationwide registry study from 2013 to 2016. ESC Heart Fail. 2020;7:1917–1926. doi: 10.1002/ehf2.12773

36. Hees PS, Fleg JL, Lakatta EG, Shapiro EP. Left ventricular remodeling with age in normal men versus women: novel insights using three-dimensional magnetic resonance imaging. Am J Cardiol. 2002;90:1231–1236. doi: 10.1016/s0002-9149(02)02840-0

37. van Essen BJ, Emmens JE, Tromp J, Ouwerkerk W, Smit MD, Geluk CA, Baumhove L, Suthahar N, Gansevoort RT, Bakker SJL, et al. Sex-specific risk factors for new-onset heart failure: the PREVEND study at 25 years. Eur Heart J. 2024. doi: 10.1093/eurheartj/ehae868

38. Rosner B. Fundamentals of biostatistics. 8th ed. Boston, Mass: Cengage Learning; 2016.

